# Proinflammatory IgG Fc structures in patients with severe COVID-19

**DOI:** 10.1101/2020.05.15.20103341

**Authors:** Saborni Chakraborty, Joseph Gonzalez, Karlie Edwards, Vamsee Mallajosyula, Anthony S. Buzzanco, Robert Sherwood, Cindy Buffone, Nimish Kathale, Susan Providenza, Markus M. Xie, Jason R. Andrews, Catherine A. Blish, Upinder Singh, Haley Dugan, Patrick C. Wilson, Tho D. Pham, Scott D. Boyd, Kari C. Nadeau, Benjamin A. Pinsky, Sheng Zhang, Matthew J. Memoli, Jeffery K. Taubenberger, Tasha Morales, Jeffrey M. Schapiro, Gene S. Tan, Prasanna Jagannathan, Taia T. Wang

**Author notes:** these authors contributed equally (Joseph Gonzalez, Karlie Edwards).

## Abstract

Severe acute respiratory syndrome coronavirus 2 (SARS-CoV-2) infections can cause Coronavirus Disease 2019 (COVID-19), which manifests with a range of severities from mild illness to life threatening pneumonia and multi-organ failure. Severe COVID-19 is characterized by an inflammatory signature including high levels of inflammatory cytokines, alveolar inflammatory infiltrates and vascular microthrombi. Here we show that severe COVID-19 patients produced a unique serologic signature, including increased IgG1 with afucosylated Fc glycans. This Fc modification on SARS-CoV-2 IgGs enhanced interactions with the activating FcγR, FcγRIIIa; when incorporated into immune complexes, Fc afucosylation enhanced production of inflammatory cytokines by monocytes, including IL-6 and TNF. These results show that disease severity in COVID-19 correlates with the presence of afucosylated IgG1, a pro-inflammatory IgG Fc modification.

---

Antibody responses to viral infections in humans are varied and of widely divergent clinical significance. Pre-existing, reactive antibodies or antibodies that are formed early during infection can bind to virus particles, forming immune complexes that can neutralize or mediate clearance of the virus. On the other hand, immune complexes can also promote inflammation and exacerbate symptoms of disease. How antibodies within immune complexes modulate infection depends, in part, on their Fc domain structure. Fc structure, in turn, dictates interactions with Fcγ receptor (FcγRs) that are expressed by a variety of cells that are activated during infection^1^.

Antibody isotypes, IgG, IgA, and IgM are a primary determinant of Fc-structure and thus of activity. Initial B cell responses are characterized by the production of IgM antibodies. Production of class-switched IgA and IgG antibodies follows, IgA playing a central role in mucosal immunity while IgG is the dominant isotype involved in systemic antiviral immunity. IgG functions are governed by interactions between immune complexes and effector immune cells that express FcγRs, the receptors for IgG. The balance of FcγRs that are engaged by immune complexes determines the degree of the inflammatory effector cell response. Activating, low-affinity FcγRs (FcγRIIa and FcγRIIIa) transduce inflammatory signaling through immunoreceptor tyrosine-based activation motifs (ITAMs); in health, ITAM signaling is balanced by immunoreceptor tyrosine-based inhibition motif (ITIM) signaling through the inhibitory FcγR, FcγRIIb^2, 3^.

The strength of interactions between immune complexes and various FcγRs is determined by structural diversity within IgG subclasses (IgG1, IgG2, IgG3, IgG4) and post-translational modifications of their Fc domains^4^. Importantly, individuals produce distinct structural repertoires of IgG Fc domains, with some producing highly activating/pro-inflammatory repertoires enriched for features such as IgG1, IgG3 and/or reduced core-fucosylation of the IgG1 Fc domain. Others produce IgG repertoires characterized by higher levels of IgG2 and/or sialylated Fcs that have reduced activating/inflammatory FcγR signaling potential^2^. It remains unknown whether there are any specific structural features of antibodies produced by mild or severe COVID-19 patients.

To address this, we performed a multi-dimensional analysis of antibodies from adult patients with severe (hospitalized) or mild (outpatient) SARS-CoV-2 infections, or from SARS-CoV-2 seropositive children, with a focus on antibody features that are known to augment effector functions. Overall, we found that severe COVID-19 was distinguished by increased production of IgG1 antibodies with significantly reduced Fc fucosylation (high afucosylation) and elevated IgG3 in patients who were treated in the ICU, both of these antibody forms being pro-inflammatory in nature. Within SARS-CoV-2 immune complexes, afucosylated Fc structures triggered activation of NK cells and production of inflammatory cytokines by primary monocytes including IL-6 and TNF.

## Results

### Severe COVID-19 patients display specific antibody signatures

43 hospitalized PCR^+^ COVID-19 patients, divided into those treated in the ICU (n=21) or on the floor (n=22), along with 18 PCR^+^ COVID-19 outpatients were studied (Extended Data Table 1). In addition, because children are rarely diagnosed with COVID-19, and almost never develop severe COVID-19 despite being susceptible to productive infections ^5, 6, 7, 8^ we reasoned that it would be informative to study SARS-CoV-2 antibodies produced by children. To this end, we screened approximately 800 remainder sera from pediatric patients in a large Northern Californian health care system and identified 16 that were positive for antibodies against the receptor binding domain (RBD) of the SARS-CoV-2 spike protein; all positive samples from children were validated in a secondary screen against the full-length spike protein, as previously described ^9^.

**Table 1.** Patient demographic data.

|  |  | ICU<br>(n=21) | Inpatient<br>Non-ICU<br>(n=22) | Outpatient<br>(n=18) | Pediatric<br>(n=16) |
| --- | --- | --- | --- | --- | --- |
| Age (median) (IQR) |  | 58<br>(41-68) | 59.5<br>(47-66.8) | 45<br>(32.8-59) | 12.5<br>(8-16.5) |
| Sex<br>(%) | Female | 7 (33.3) | 8 (36.3) | 10 (55.6) | 8 (50) |
|  | Male | 14 (66.7) | 14 (63.7) | 8(44.4) | 8 (50) |

To characterize serologic correlates of disease severity, we first profiled anti-RBD immunoglobulin isotype titers in sera from COVID-19 patients or from seropositive children. Among the study subjects, severe COVID-19 patients (ICU and floor) had significantly higher serum titers of IgM and IgA RBD-binding antibodies compared to both mild COVID patients and seropositive children. The titers of anti-RBD IgG antibodies were not significantly different amongst the groups (Fig.1a, 1b).

**Figure 1.**
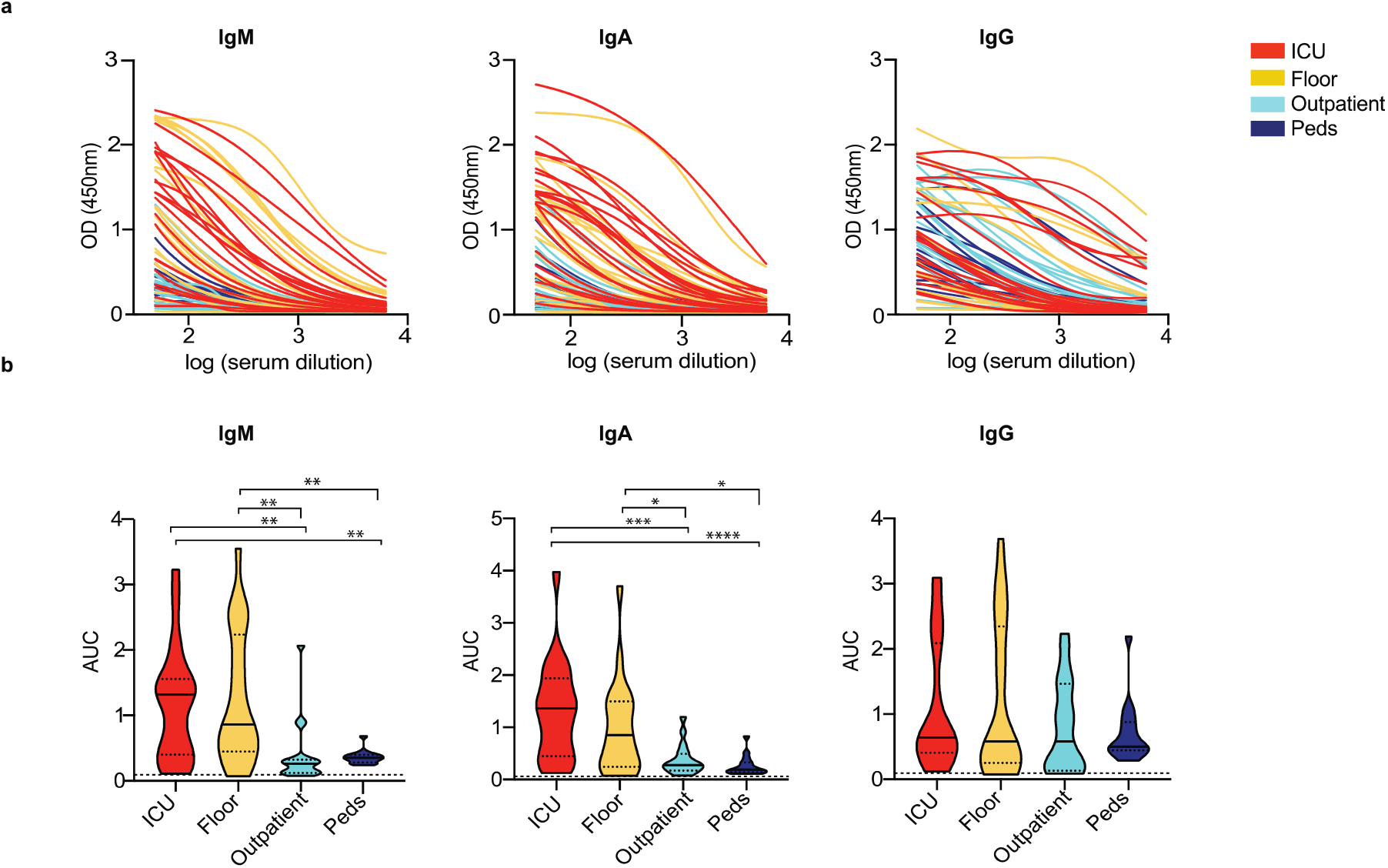
SARS-CoV-2 antibodies in COVID-19 patients and in undiagnosed children. **(a)** Anti-RBD IgM, IgA, IgG titers in COVID patients who required treatment in the ICU (red) (n= 21), hospitalization but no ICU (Floor, yellow) (n=22), patients treated on an outpatient basis (Outpatient, light blue) (n=18), or seropositive children (Peds, dark blue) (n=16) (b) Anti-RBD AUC for the four patient groups are shown. Data shown are representative of at least two experiments performed in triplicate. Violin plots show the distribution of sample values along with median (solid lines) and quartile (broken lines) values. The dashed line indicates the average AUC value of the pre-COVID-19 historical samples. P values in (b) were calculated using one-way ANOVA with Tukey’s multiple comparisons testing between all groups. (IgM- p=0.0079 ICU vs. Outpatient, p=0.0079 ICU vs Peds, p=0.0023 Floor vs Outpatient, p=0.0023 Floor vs Peds. IgA- p=0.0005 ICU vs. Outpatient, p=0.0001 ICU vs Peds, p=0.0454 Floor vs Outpatient, p=0.0148 Floor vs Peds.). *P ≤ 0.05, **P ≤ 0.01, ***P ≤ 0.001, ****P ≤0.0001.

Next, we defined the determinants of anti-RBD IgG effector function in the COVID-19 patients and seropositive children. The absolute abundance of IgG subclasses was characterized by mass spectrometry. We observed that the abundance of various subclasses was similar between groups, with a small but significant increase in IgG3 produced by COVID-19 patients who were in the ICU (Fig. 2a). Anti-RBD IgG1, the most abundant IgG subclass, was next characterized for post-translational modifications of the Fc using well-established mass spectrometric methods ^10, 11, 12^. The relative abundance of N-glycan modifications were characterized, including fucose, bisecting GlcNAc, galactose and sialic acid. Notably, anti-RBD IgG1 from severe COVID-19 patients was significantly reduced in core fucosylation when compared with anti-RBD IgG1 from mild COVID-19 patients or from seropositive children (Fig. 2b, 2c). We validated the ability of reduced fucosylation to separate patients with severe COVID-19 from those with mild disease using receiver operating curve (ROC) analysis (global area under the ROC curve (AUC) 0.74 [95% CI 0.62–0.86], p= 0.0031) (Fig. 2d). Of all post-translational modifications, IgG1 Fc fucosylation was a uniquely strong marker of severe COVID-19 using ROC analysis (Extended Data Fig. 1). When we analyzed the six afucosylated Fc glycoforms that were quantified, those without a bisecting N-acetyl glucosamine (F0N0) were specifically increased in severe COVID-19 (Fig. 2e).

**Figure 2.**
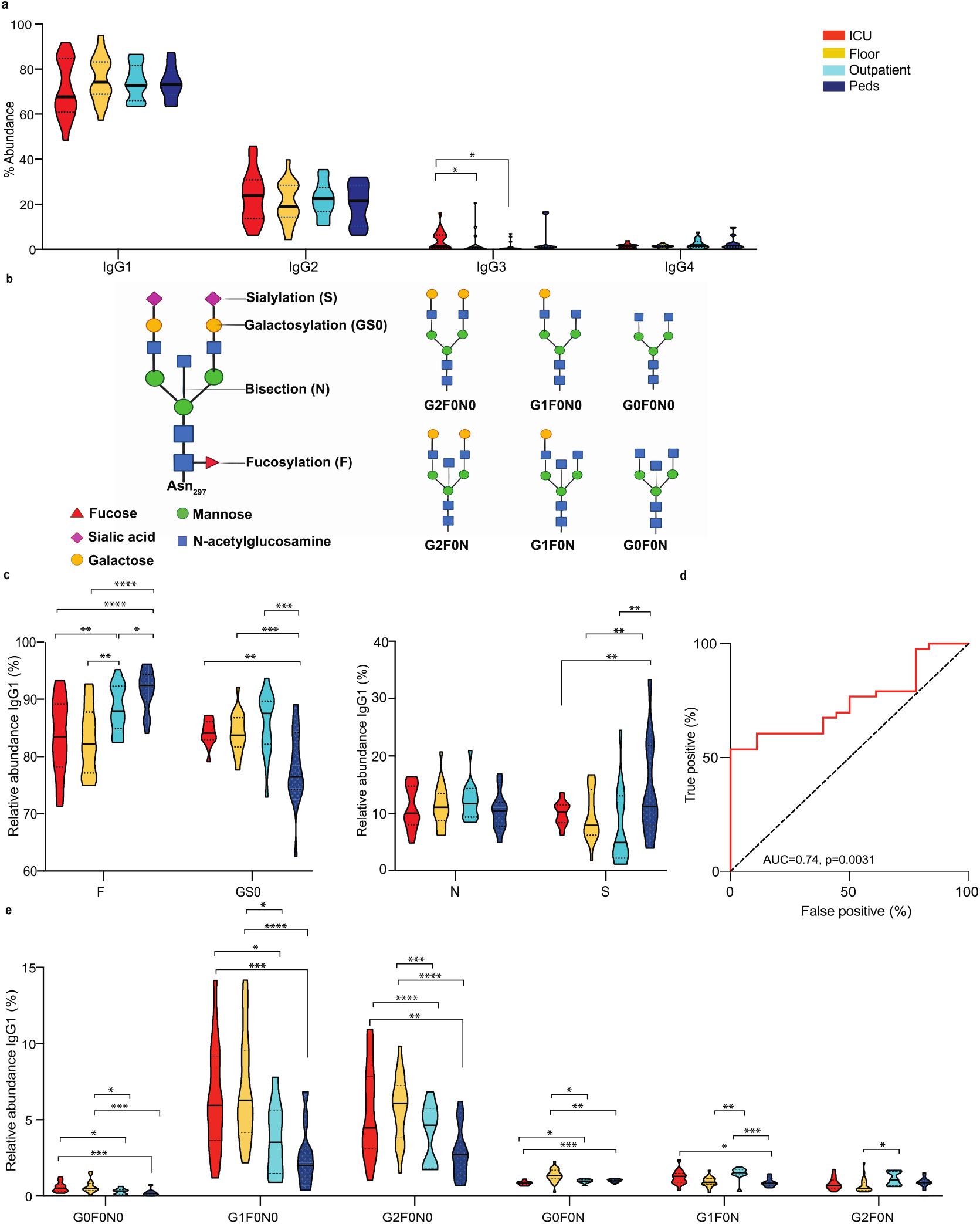
Structural properties of anti-RBD IgG Fc domains in adult COVID-19 patients and in seropositive children. (a) The abundance of anti-RBD IgG subclasses was characterized. ICU patients had elevated levels of IgG3 compared to those treated on the floor or outpatients. P values were calculated using unpaired two-tailed t-tests (p=0.0449 for ICU vs Floor, p=0.0243 for ICU vs Outpatient). (b) Cartoon representation of IgG1 Fc glycans and various F0 modifications that were characterized. (c) Anti-RBD IgG1 Fc post-translational modifications were characterized. Patients who were hospitalized (ICU or floor) had significantly reduced Fc fucosylation (F), when compared with RBD IgGs from outpatients (p=0.0092 for ICU vs Outpatient and p=0.0011 for Floor vs Outpatient) or from children (p=0.0001 ICU vs Peds and p=<0.0001 for Floor vs Peds). Fc galactosylation (GS0) was significantly higher and sialylation (S) significantly lower in all adult patients compared with children (GS0: p=0.0019 ICU vs Peds, p=0.0007 for Floor vs Peds and p=0.0007 for Outpatient vs Peds. S: p=0.0097 for ICU vs Peds, p=0.0045 for Floor vs Peds and p=0.0052 for Outpatient vs Peds). No significant differences were observed in levels of IgG1 Fc bisection (N). (d) Receiver operating characteristic (ROC) curve for anti-RBD IgG1 fucosylation from all hospitalized (n=43) and mild (n=18) COVID-19 patients showed IgG1 fucose levels separated the two cohorts. Area under the curve (AUC) 0.74 [95% CI (0.62-0.86, p=0.0031). (e) Of the six afucosylated forms quantified, those lacking both core fucose and a bisecting N-acetyl glucosamine (F0N0) were substantially enriched in severe COVID-19 patients. Violin plots show the distribution of sample values along with median (solid lines) and quartile (broken lines) values. P values in (c) and (e) were calculated using one-way ANOVA with Tukey’s multiple comparisons testing between all groups. *P ≤ 0.05, **P ≤0.01, ***P ≤ 0.001, ****P≤ 0.0001

Overall, these data showed a unique antibody signature associated with COVID-19 severity based on multiparametric characterization of SARS-CoV2-specific humoral responses comprising isotype titers, IgG subclasses and IgG1 Fc-glycan structures. Severe COVID-19 patients were more likely to produce significantly higher titers of anti-RBD IgM and IgA isotype antibodies, increased IgG3 subclass (ICU patients) and increased afucosylated (F0N0) Fc glycoforms relative to patients with mild COVID-19 (Fig. 3). The IgG3 subclass and afucosylated IgG1 modifications are features that increase Fc interactions with activating/pro-inflammatory FcγRs ^2^.

**Figure 3.**
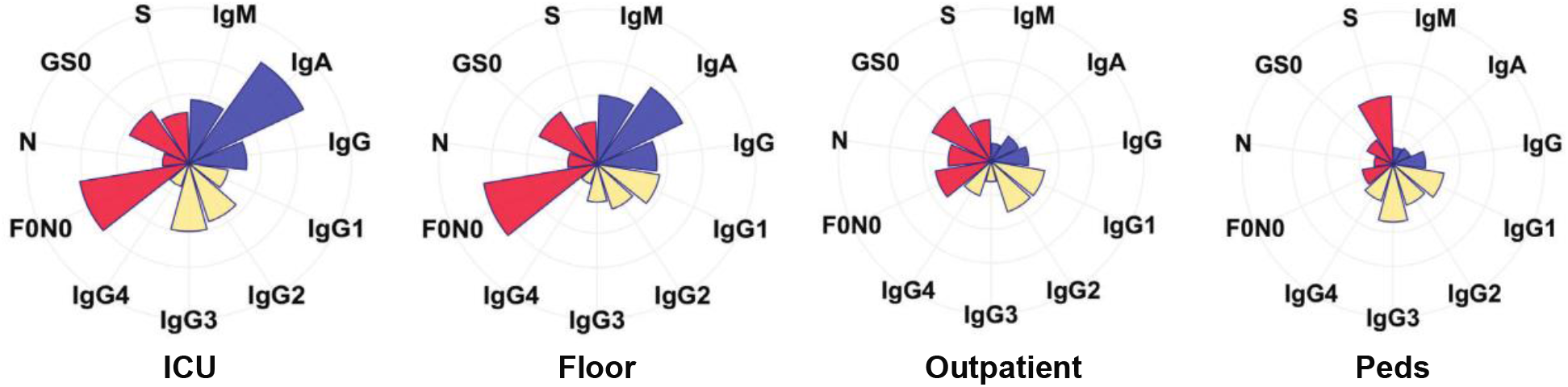
Summary of antibody signatures from COVID-19 patients. Relative multi-dimensional antibody signatures for each group stratified by disease severity are depicted by radar plots. Each feature, isotype (IgM, IgA, IgG) in blue/purple, IgG subclass (IgG1, IgG2, IgG3 and IgG4) in yellow and % abundance of IgG1 Fc glycoforms (F0N0, GS0, S and N) in red/pink, is depicted as a wedge. The size of the wedges indicates the median of the features, normalized to the corresponding outpatient feature.

### Fc afucosylation is stable over time in severe COVID-19

The afucosylated IgG1 Fc was the most prominent feature distinguishing severe and mild COVID-19 in our cohorts. To begin to dissect the regulation of this antibody modification we first asked whether infection itself triggered a significant change in Fc glycoforms. To do this, we performed a longitudinal analysis of Fc glycoforms on RBD-reactive IgG1 from paired sera drawn at acute (T1) and later time points. Paired samples were selected from individuals who were positive for IgG at T1 and late time points were drawn 2, 3- or 4-weeks post T1. No significant changes were found in the levels of F0N0, bisection or galactosylation over time but the amount of sialylation was significantly reduced between T1 and week-4 samples (Supplementary Figure 4a). This suggested that infection triggered an acute increase in Fc sialylation that declined over time as new IgGs were produced but this was not the case for other glycoforms. In contrast to SARS-CoV-2 infection, it was previously observed that acute dengue virus infections can trigger production of highly afucosylated IgGs that decline in the weeks following infection^12^. To further probe whether anti-RBD Fc glycoforms were associated with SARS-CoV-2 viral load, we performed correlation analyses between anti-RBD Fc glycoforms and SARS-CoV-2 RNA levels from nasopharyngeal swabs taken during acute infection. We observed no correlation between any Fc glycoforms and viral RNA load as determined by the cycle threshold (CT) value (Supplementary Figure 4b). In all, these data did not support the hypothesis that SARS-CoV-2 virus infection regulated F0N0 Fc glycans during infection.

**Figure 4.**
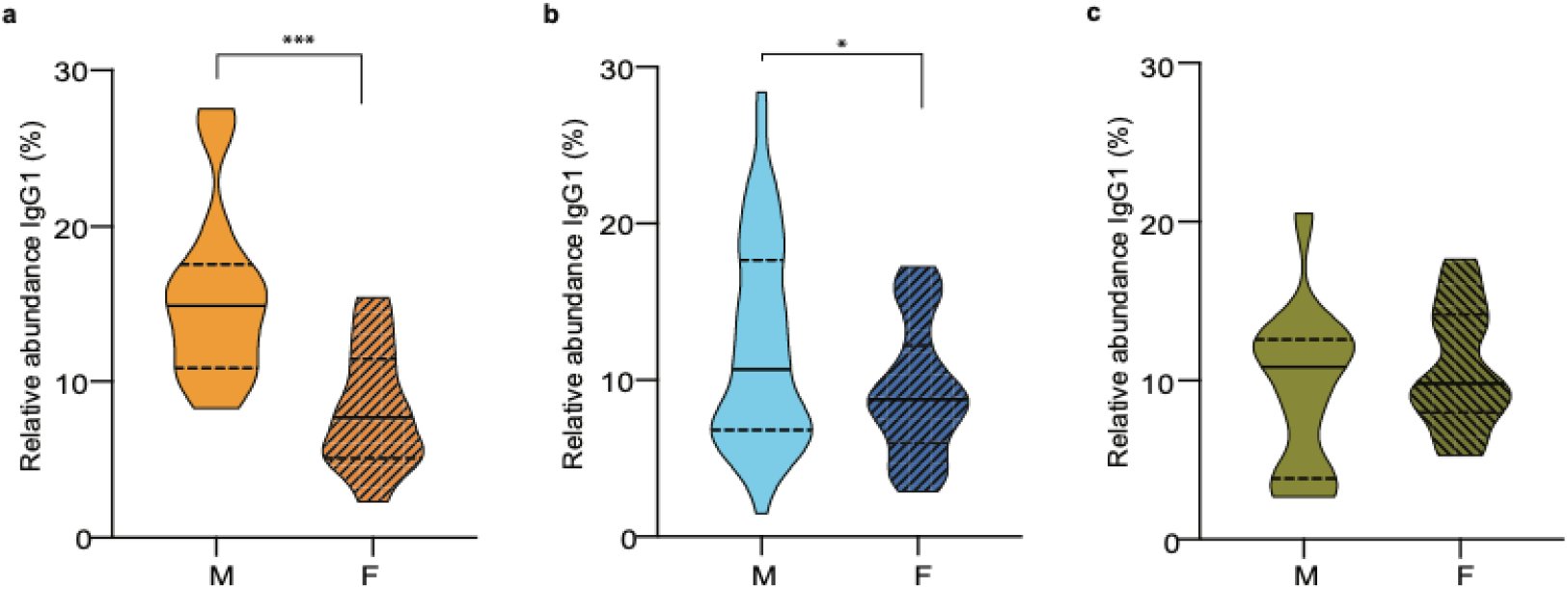
anti-SARS-CoV2 IgG1 Fc afucosylation varies by sex. The level of the afucosylated anti-RBD IgG1 was significantly higher in hospitalized males, compared to hospitalized females in two different cohorts from (a) Stanford Hospital Center (n=30, F=14, M=16) (p=0.0007) or (b) Kaiser Permanente Hospitals of Northern California (n=81, M=55, F=26) (p=0.0190). (c) Significant sex associated differences in afucosylation levels of IgG1 were not present in mild COVID-19 (outpatients) (n=27, F=14, M=13). Violin plots in (a), (b) and (c) show the distribution of sample values along with median (solid lines) and quartile (broken lines) values. P values were calculated using a two-tailed unpaired t test with Welch’s correction. *P ≤ 0.05, **P ≤ 0.01, ***P ≤ 0.001, ****P ≤ 0.0001

### Fc afucosylation is elevated in males

Studies have previously shown that antibody glycans may be influenced by age and sex^13, 14, 15, 16^. We therefore performed a multivariate regression analysis on samples from hospitalized COVID-19 patients from the Stanford Hospital Center (n=30, F=14, M=16) to determine whether age and/or sex may have played a role in regulating the abundance of Fc glycans. We observed that sex was significantly correlated with Fc afucosylation (p=0.0007) and age was not a confounding variable. Males had significantly higher levels of anti-RBD F0N0 Fc glycoforms over females (Figure 4a). To determine the generalizability of this finding, we studied samples from a second cohort of hospitalized COVID-19 patients treated in Northern California Kaiser Permanente hospitals (n=81, M=55, F=26). In this larger cohort, males also had significantly elevated anti-RBD F0N0 Fc glycoforms over females (p=0.019) (Figure 4b). A similar sex associated difference in levels of afucosylation was not observed in mild COVID-19 patients (n=27, F=14, M=13) (Figure 4c). No other Fc glycoforms segregated by sex in any of the cohorts (Extended Figures 4c-e). Further, age did not correlate with any of the characterized anti-RBD Fc glycoforms (Extended Figure 5). Overall, these data showed that males with severe COVID-19 produced higher levels of afucosylated, anti-RBD IgG1 antibodies over females.

**Figure 5.**
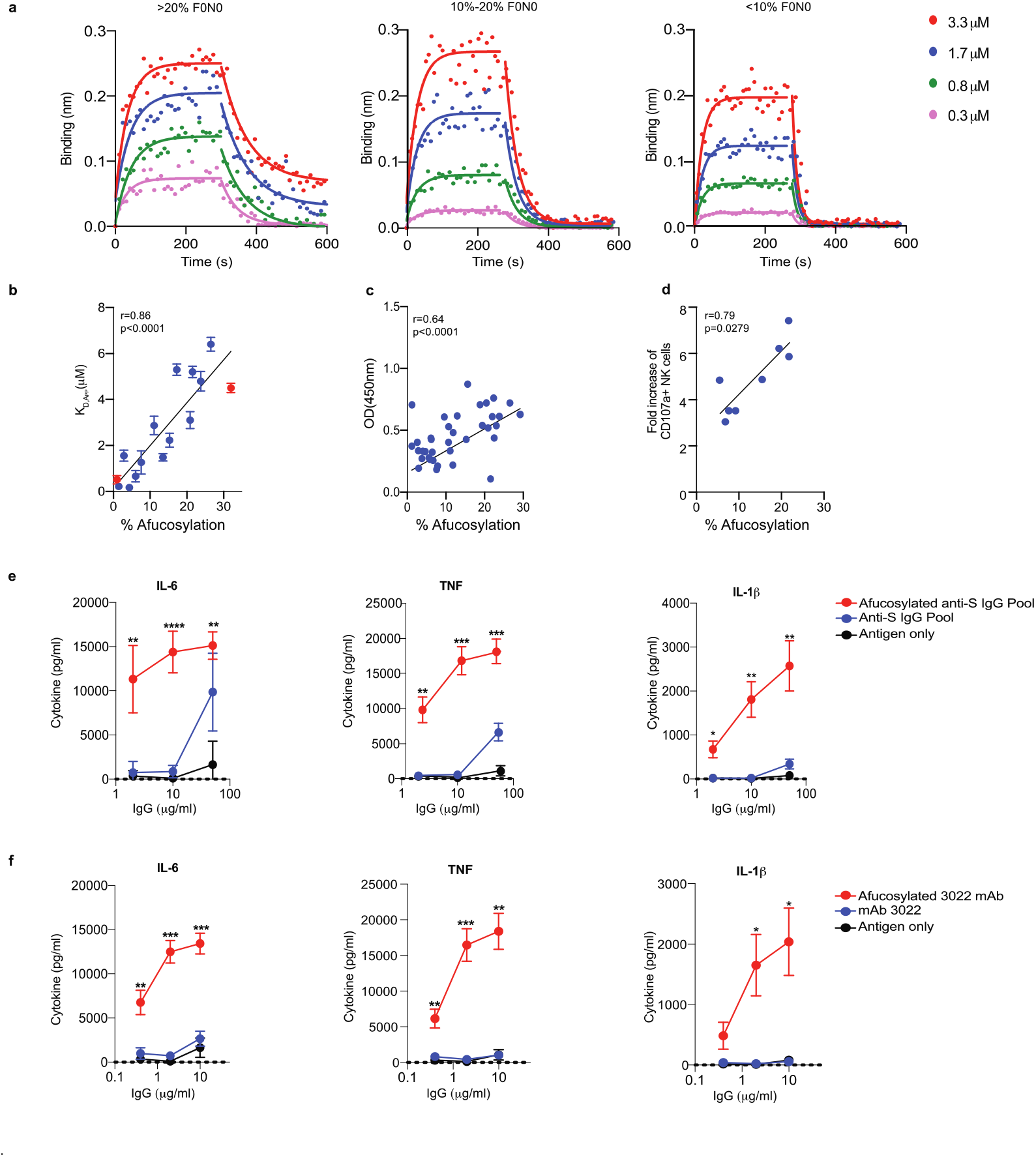
FcγRIIIa binding by human anti-RBD IgGs with variable Fc fucosylation. **(a)** Binding of polyclonal IgGs was determined by biolayer interferometry. The overlay of binding traces for a donor from each group representing varying degree of anti-RBD core afucosylation; low (0-10%), medium (10-20%) and high (>20%) is shown. The kinetic constants were obtained by evaluating the binding at multiple concentrations (3.3 mM followed by 2-fold dilutions) of the analyte as shown (solid circles). The fits are indicated by solid lines. The assay was performed twice and shown are representative traces from one experiment. (b) A strong positive correlation (Pearson correlation coefficient r=0.86) was observed between the apparent dissociation constant (KD,app) of FcγRIIIa (CD16a) and anti-RBD IgG1 core afucosylation in COVID-19 patients (n=13) as determined by biolayer interferometry. The binding of anti-RBD monoclonal antibody, CR3022 with different core afucosylation levels is also shown (red). The mean data and the standard error of the mean (SEM) have been graphed. (c) Correlation between the level of anti-RBD IgG1 Fc afucosylation and binding to FcγRIIIa. Binding of serum antibodies (n=38) to FcγRIIIa correlated positively with the degree of afucosylation (Pearson correlation coefficient r=0.64). Samples were representative of the range of Fc fucosylation over the sample set. (d) Correlation between the level of anti-RBD IgG1 Fc afucosylation and immune complex (IC) mediated NK cell degranulation. The amount of degranulation measured by fold increase of CD107a+ NK cells over control correlated positively with the degree of afucosylation of the anti-RBD IgG (Spearman correlation coefficient r= 0.79). The assay was performed in duplicate with PBMCs from three healthy donors and mean data has been graphed. (e, f) Highly afucosylated immune complexes elicited increased production of inflammatory cytokines IL-6, TNF, and IL-1b. Immune complexes were formed using (e) pooled polyclonal IgGs from COVID patients (IL6: p=0.0081 for 100µg/ml, p <0.0001 for 20µg/ml and p=0.0014 for 4µg/ml. TNF: p= 0.0042 for 100µg/ml, p=0.0004 for 20µg/ml and p= 0.0042 for 4µg/ml. IL-1β: p=0.0068 for 100µg/ml, p=0.0065 for 20µg/ml and p=0.0198 for 4µg/ml) or (f) recombinant IgG1 mAb 3022 with distinct levels of afucosylation) (IL6: p=0.0009 for 10µg/ml, p=0.0001 for 2µg/ml and p=0.0017 for 0.4µg/ml. TNF: p= 0.0022 for 10µg/ml, p=0.0008 for 2µg/ml and p= 0.0022 for 0.4µg/ml. IL-1β: p=0.0152 for 10µg/ml, p=0.0231 for 2µg/ml). The assays were performed in duplicate with monocytes from three healthy donors and mean data and standard error of the mean (SEM) has been graphed. P-values between high and low afucosylated immune complexes at each antibody concentration were calculated by two-tailed paired t-tests. *P ≤ 0.05, **P ≤0.01, ***P ≤ 0.001, ****P ≤ 0.0001

### Afucosylated SARS-CoV-2 immune complexes can promote FcγRIIIa interactions and inflammatory cytokine production

Afucosylation of IgG1 Fc confers 5-10-fold higher affinity for the activating FcγR, FcγRIIIa on a monomeric basis (and higher affinity in the context of a multivalent complex), relative to fucosylated IgG1 ^17^. FcγRIIIa is present on monocytes, macrophages and NK cells at baseline and can enhance cell activation, pro-inflammatory cytokine production and cytotoxic effector cell activity ^18, 19^. Thus, distinct levels of fucosylation within SARS-CoV-2 immune complexes would be expected to modulate their binding to FcγRIIIa and activating ITAM signaling^17^. To determine whether serum IgGs from study subjects differed in their capacity to bind FcγRIIIa, we measured the apparent dissociation constant of purified serum IgGs from severe COVID-19 patients with a range of fucose levels, to recombinant FcγRIIIa (F158). IgGs from patients with RBD IgG1 afucosylation >20% conferred ∼3-fold higher affinity to FcγRIIIa over IgGs with medium afucosylation and 5-6 fold over IgGs with low IgG1 afucosylation (<10%). Further, RBD IgG1 afucosylation levels correlated with the apparent dissociation constant (Kd) of IgGs for FcγRIIIa (p<0.0001)(Figures 5a, 5b). To further characterize this using homogenous Fc structures, we expressed the anti-SARS-CoV2 RBD monoclonal antibody (mAb) CR3022^20, 21^ as a fully fucosylated IgG1 or as a variant with 32.6% afucosylated Fc, as determined by mass spectrometry; these variants differed in apparent kD for FcγRIIIa by ∼7-fold (Figure 5b). Next, we performed an ELISA to measure binding by IgGs from individual sera to FcγRIIIa. 38 sera were selected that represented the range of afucosylation over the sample set. Consistent with the binding results using purified IgGs, we observed that the amount of anti-RBD IgG1 afucosylation correlated with binding to FcγRIIIa (p<0.0001) (Figure 5c).

To determine whether these binding differences were physiologically relevant to activation of primary immune cells that express FcγRIIIa, we next performed in vitro stimulation assays. We first assessed activation of NK cells by immune complexes composed of patient IgGs and RBD. NK cell degranulation, measured by CD107a-positive staining, correlated with the abundance of anti-RBD afucosylation in immune complexes (p=0.0279) (Figure 5d). We next measured the ability of immune complexes with different levels of Fc afucosylation to stimulate cytokine production by primary monocytes. Immune complexes were formed from pooled IgGs mixed with SARS-CoV-2 pseudoparticles; these complexes were added to primary cells from separate donors. IgG pools were derived from patients with high (>20%) or low (<10%) anti-RBD IgG1 afucosylation and were matched for SARS-CoV-2 IgG binding titer (Supplementary Figure 6). Highly afucosylated immune complexes triggered increased production of proinflammatory cytokines, principally IL-6, TNF and IL-1β compared to immune complexes with low afucosylation (Figure 5e, Supplementary Figures 7a). To better define the role of Fc fucosylation alone on the differential activation of monocytes, we performed this assay with immune complexes made from SARS-CoV-2 pseudoparticles along with anti-RBD mAb 3022 variants that differed only in Fc fucosylation, as described above. This modification had no impact on anti-spike binding (Supplementary Figure 6). Cytokines including IL-6, TNF and IL-1β were significantly enhanced after incubation of monocytes with afucosylated mAb 3022 complexes (Figure 5f, Supplementary Figures 7b).

Overall, differences in core IgG1 afucosylation impacted FcγRIIIa binding and, within SARS-CoV-2 immune complexes, enrichment of afucosylated Fc structures could promote production of cytokines including IL-6, TNF and IL-1β by primary monocytes. Thus, severe COVID-19 patients were more likely to produce a pro-inflammatory form of IgG antibody (Extended Data Fig. 6).

## Discussion

These studies show that specific pro-inflammatory antibody forms, characterized by IgG3 and IgG1 with F0N0 glycoform modification are elevated in more people with severe COVID-19. This was in contrast to patients with mild symptoms and seropositive children. We also found that male sex, in particular, was associated with high Fc afucosylation in severe COVID-19. Because prior studies in healthy adults have not found a sex-associated variation in IgG1 afucosylation, this correlation may arise from additional variables that were not identified in this study^13, 14, 15^.

Low affinity activating FcγR pathways (FcγRIIa and FcγRIIIa) are often required for potent immunity mediated by passively transferred antibodies against viral pathogens^22^ and, indeed, one or both of these receptors likely has a role in protective antibody functions against SARS-CoV-2. The balance of activating to inhibitory FcγR signaling that promotes protective immunity and the thresholds for activating FcγR signals that may promote inflammatory pathology will be an important topic for future investigations into immunity against SARS-CoV-2. Overall, the data shown here raise the possibility that immune complex-mediated activation of inflammatory FcγR pathways and the associated cytokine production is one “hit” that can promote progression to severe COVID-19. A limitation of the present study is the absence of *in vivo* experimental data to define this due to the current lack of a humanized FcγR animal model for COVID-19 that would enable such mechanistic studies. *In vivo* models with well characterized FcγR-IgG interactions and signaling outcomes will be essential for defining the role of antibodies in SARS-CoV-2 immunity and disease. Interestingly, prior studies have found high levels of afucosylated IgG1 in patients with severe dengue virus infections and elevated maternal anti-dengue afucosylation predicted risk for dengue disease in their infants^10, 12^. Mechanistic studies have shown that afucosylated anti-dengue immune complexes and the associated enhancement in FcγRIIIa ITAM signaling can modulate diverse aspects of dengue virus-host interactions^10, 12^.

These studies show that production of pro-inflammatory IgG antibodies was more common in severe cases of COVID-19. Future longitudinal studies including analysis of pre- and post-infection samples will be needed to determine whether these Fc structures can be used as a pre-infection biomarker for risk of progression to severe COVID-19.

## Data Availability

All raw data are available from the corresponding author on reasonable request.

## Acknowledgments

Support was received from Stanford University, the Chan Zuckerberg Biohub and the Searle Scholars Program. Research reported in this publication was supported by Fast Grants, CEND COVID Catalyst Fund, the National Institute Of Allergy And Infectious Diseases of the National Institutes of Health under Award Numbers U19AI111825, U54CA260517 and R01AI139119, R01AI130398 and R01AI127877. Clinical samples were obtained with support from the Rockefeller University Center for Clinical and Translational Science Grant # UL1 TR001866. The content is solely the responsibility of the authors and does not necessarily represent the official views of the National Institutes of Health. TTW thanks C. Kopel for exceptional technical support.

## Author contributions

T.T.W, S.C., K.E., J.G. conceived of and/or designed experiments. T.T.W, S.C., K.E., R.S., S.Z., V.M., A.S.B., M.M.X., J.G., C.B., N.K., S.P., J.R.A., J.M.S, T.M., C.A.B., P.C.W, T.D.P., S.D.B., U.S., B.A.P., K.C.N., H.D., M.J.M, J.K.T., G.S.T., P.J. were involved in data acquisition and/or analysis and/or interpretation. S.C. and T.T.W wrote the manuscript.

## Methods

### Cloning, expression and protein purification

The His_6_-tagged SARS-CoV-2 RBD construct was a kind gift from F. Krammer (Icahn School of Medicine at Mount Sinai). The full length recombinant SARS-CoV-2 spike protein (residues 1−1208 (GenBank:MN908947, protein id QHD43416.1)) construct was designed with the following modifications: two well-characterized proline substitutions (K986P and V987P) ^23^; a four amino acid substitution to remove the furin cleavage site (RRAR → GSAS) in order to stabilize the pre-fusion conformation ^24^; a synthetic trimerization motif-the globular β-rich ‘foldon’ from T4 fibritin to promote oligomerization in lieu of the native trans-membrane (TM) domain; a human rhino virus 3C (HRV 3C) protease cleavage site; a C-terminal His_8_ tag. Mammalian codon-optimized gene fragments were synthesized (Integrated DNA Technologies, Inc.) and cloned using Gibson Assembly (New England BioLabs) into a CMV/R promoter driven mammalian expression vector between XbaI and BamHI restriction sites.

Both the constructs were transiently transfected into Expi293F cells (Thermo Fisher Scientific) as per the manufacturer’s recommendations. Briefly, Expi293F cells at a density of 3×10^6^ viable cells/ml maintained in Expi293 expression medium (Thermo Fisher Scientific) were transfected with expression plasmids complexed with ExpiFectamine 293 transfection reagent. 18 hours post-transfection, the cells were supplemented with a cocktail of transfection enhancers. The cultures were incubated for four days, following which the culture supernatants were harvested by centrifugation for protein purification. The supernatants were incubated with phosphate-buffered saline (PBS (Gibco))-equilibrated Ni-nitriloacedic acid (NTA) resin (GE HealthCare) for 2h at 4°C under mild-mixing conditions to facilitate binding. The proteins were subsequently eluted using 500mM imidazole (in PBS, pH 7.4) under gravity flow. The eluted fractions were pooled, buffer exchanged into PBS (pH 7.4) and concentrated using Amicon Ultra centrifugal units (EMD Millipore) to a final concentration of ∼1mg/ml. Protein purity was assessed by sodium dodecyl sulfate–polyacrylamide gel electrophoresis (SDS-PAGE). Size exclusion chromatography was used to determine the oligomeric state of the purified proteins under non-denaturing conditions at room temperature on a Superdex-200 analytical gel filtration column and data was acquired on UNICORN 7 software (Cytiva). For molecular weight estimations, the column was calibrated using broad range molecular weight markers (Cytiva) (Figure S1).

MAb 3022^21^ was a kind gift from I. Wilson (The Scripps Research Institute) and was produced in Expi293F cells, as described above. For production of afucosylated mAb 3022, 200μM of fucosyl transferase inhibitor 2F-Peracetyl-Fucose (Sigma Aldrich) was added to the culture after transfection. Supernatants were harvested 5 days after transfection and antibody purifications were done over Protein G Sepharose 4 Fast Flow resin (GE Healthcare). Antibodies were buffer-exchanged into PBS pH 7.4 and concentrated using Amicon Ultra centrifugal units (EMD Millipore).

### ELISAs

#### Screening ELISA

A rapid, high-throughput screening ELISA was performed on a total of 789 pediatric samples to test seropositivity following a modified version of a protocol described previously. Briefly, round bottom 96 well plates (Immunolon 2HB (Thermo Scientific)) plates were coated with 50μl of RBD at 2μg/ml in PBS for 1h at room temperature (RT). Next, the plates were blocked for an hour with 3% non-fat milk in PBS with 0.1% Tween 20 (PBST). All serum samples from COVID-19 patients, the pediatric cohort and the negative controls were heated at 56°C for 1h, aliquoted and stored at −80°C. For the first round of screening, all samples were diluted 1:50 in 1% non-fat milk in PBST. 50μl of the diluted sera was added to each well and incubated for 2h at RT. Following primary incubation with the sera, 50μl of 1:5000 diluted horse radish peroxidase (HRP) conjugated goat anti-Human Ig Fab (Southern Biotech) was added and incubated for 1h at RT. The plates were developed by adding 50ul/well of the chromogenic substrate 3,3′,5,5′-tetramethylbenzidine (TMB) solution (Millipore Sigma). The reaction was stopped with 0.2N sulphuric acid (Sigma) and absorbance was measured at 450nm (SPECTRAmax 250, Molecular Devices). The plates were washed 5 times with PBST between each step and an additional wash with PBS was done before developing the plates. Samples were considered seropositive against RBD if their absorbance value was greater than the mean plus four standard deviation (SD) of all negative controls (n=130).

### Validation ELISA of seropositive pediatric samples

The serum samples from the pediatric cohort, which showed seropositivity against RBD were validated by a second round of screening against the full-length SARS-Cov-2 spike protein (S). As described above, plates were coated with 50ul of 2ug/ml S protein in PBS. Following blocking and wash, the plates were incubated with a 5-fold dilution series of RBD positive sera (50μl) starting at 1:50 for 2h at room temperature. All the subsequent steps were followed as described above. Sera from children and COVID-19 patients were considered positive if they reached a threshold of the average value of 130 historical negative controls plus six standard deviations. The specificity of the assay was also tested on control sera from 12 subjects with documented seasonal coronavirus infections collected in early 2019.

### Isotyping by ELISA

Sera were diluted 5-fold starting at 1:50 and ELISAs were performed as described above. The various secondary antibodies used for isotyping were 1:5000 dilutions of HRP-conjugated Goat Anti-Human IgG Fc (Southern Biotech), Mouse Anti-Human IgM (Southern Biotech) and Goat Anti-Human IgA Fc (Southern Biotech).

### CD16a ELISA

Human recombinant CD16a (Sino Biological) was immobilized at 3ug/ml (50ul/well) in PBS at 4°C overnight, followed by an hour of blocking with 3% non-fat milk in PBST. 50ul of 1:50 diluted sera from pediatric seropositive (n=11), PCR^+^ and seropositive (n=22) and healthy controls (n=5) were added to each well and incubated for 2h at 37°C. Subsequently the plates were incubated for 1h at 37°C with 1:5000 dilution of HRP-conjugated Goat Anti-Human IgG Fc (Southern Biotech) secondary antibody, developed as described previously with TMB and absorbance was recorded at 450nm.

### Clinical cohorts and samples

Remnant sera from pediatric subjects and from PCR^+^ COVID-19 patients were obtained from Kaiser Permanente Northern California. The sera were collected for a variety of clinical tests at one of 75 distinct hospitals or outpatient clinics across 17 counties in Northern California between March 30^th^, 2020 to April 19^th^, 2020. Additional serum samples from PCR^+^ COVID-19 patients were from the Stanford ICU Biobank (protocol #28205) or from (protocol #NCT04331899) ^25^. Characterization of these samples was performed under a protocol approved by the Institutional Review Board of Stanford University (protocol #55718).

Samples from people with seasonal coronavirus infections were collected at the University of Chicago. Samples were de-identified serums of healthcare workers that had respiratory illnesses, were swabbed, and tested positive for common cold Corona virus infections in 2019 (U. Chicago protocol # 09-043-A).

Historical controls and healthy controls: 30 samples from a US cohort was enrolled at the Rockefeller University Hospital in New York City in 2012 in accordance with a protocol approved by the Institutional Review Board of Rockefeller University (protocol #TWA-0804), in compliance with guidelines of the International Conference on Harmonization Good Clinical Practice guidelines, and was registered on www.clinicaltrials.gov (NCT01967238). Blood samples were drawn from healthy adult volunteers between the ages of 18-64. 50 samples were obtained from a Ugandan cohort of women and children enrolled in PROMOTE (NCT 02163447), a randomized clinical trial of novel antimalarial chemoprevention regimens in Eastern Uganda ^26^. The study was approved by the Institutional Review Boards of the Makerere University School of Biomedical Sciences, the Uganda National Council for Science and Technology, and the University of California San Francisco. Written informed consent was obtained from all study participants. 50 samples were obtained from children under 18 years of age enrolled in a study of acute febrile illness in Nepal. The study was approved by the Nepal Health Research Council, Kathmandu University Institutional Review Board, and Stanford University Institutional Review Board (protocol #29992).

### IgG Fc glycan and IgG subclass analysis

Methods for relative quantification of Fc Glycans and IgG subclasses have been previously described^11, 12^. Briefly, IgGs were isolated from serum by protein G purification. Antigen-specific IgGs were isolated on NHS agarose resin (ThermoFisher; 26196) coupled to the protein of interest. Following tryptic digestion of purified IgG bound to antigen-coated beads, nanoLC-MS/MS analysis for characterization of glycosylation sites was performed on an UltiMate3000 nanoLC (Dionex) coupled with a hybrid triple quadrupole linear ion trap mass spectrometer, the 4000 Q Trap (SCIEX). MS data acquisition was performed using Analyst 1.6.1 software (SCIEX) for precursor ion scan triggered information dependent acquisition (IDA) analysis for initial discovery-based identification.

For quantitative analysis of the glycoforms at the N297 site of IgG1, multiple-reaction monitoring (MRM) analysis for selected target glycopeptide was applied using the nanoLC-4000 Q Trap platform to the samples which had been digested with trypsin. The m/z of 4-charged ions for all different glycoforms as Q1 and the fragment ion at m/z 366.1 as Q3 for each of transition pairs were used for MRM assays. A native IgGs tryptic peptide (131-GTLVTVSSASTK-142) with transition pair of, 575.9^+2^/780.4 was used as a reference peptide for normalization. IgG subclass distribution was quantitatively determined by nanoLC-MRM analysis of tryptic peptides following removal of glycans from purified IgGs with PNGase F. Here the m/z value of fragment ions for monitoring transition pairs was always larger than that of their precursor ions to enhance the selectivity for unmodified targeted peptides and the reference peptide. All raw MRM data was processed using MultiQuant 2.1.1 (SCIEX). All MRM peak areas were automatically integrated and inspected manually. In the case where the automatic peak integration by MultiQuant failed, manual integration was performed using the MultiQuant software.

### Binding affinity measurements using biolayer interferometry (BLI)

Total IgGs were purified from sera of 13 COVID-19 patients Protein G beads. The binding affinities of patient IgGs and high and low afucosylated 3022 mAbs were determined by biolayer interferometry (BLI) using an OctetQK instrument (Pall ForteBio). Human recombinant CD16a (Sino Biological) was captured onto the amine reactive second-generation (AR2G) biosensors using the amine reactive second-generation reagent kit (Pall ForteBio). The CD16a bound sensors were dipped into a concentration series (3.33μM, 1.7μM, 0.832μM and 0.33μM) of IgGs in PBST to determine the binding kinetics. An equal number of unliganded biosensors dipped into the analytes served as controls for referencing. The traces were processed using ForteBio Data Analysis Software version 8.0.3.5 and corrected for non-specific binding. The data was fitted globally to a simple 1:1 Langmuir interaction model.

### NK cell degranulation assay

PBMCs were isolated from whole blood collected from healthy blood donors post-plateletpheresis (Stanford Blood Center) using SepMate Isolation Tubes (STEMCELL). Cells were plated in a 96-well round-bottom plate (CELLSTAR) at a density of 3×10^6^ cells/mL of complete RPMI-1640 media supplemented with 1X penicillin-streptomycin-glutamine, 1mM sodium pyruvate, and 1X MEM Non-Essential Amino Acids, 10% heat-inactivated fetal bovine serum (Gibco), and 1ng/mL IL-15 (STEMCELL) and rested overnight at 37°C in a 5% CO_2_ incubator (Panasonic). The following morning, cell culture media was replaced with complete RPMI containing anti-CD107a antibody (BioLegend; clone H4A3). PBMCs were promptly stimulated for 6hr at 37°C with immune complexes formed by incubating purified patient IgG with SARS-CoV-2 receptor-binding domain protein at a molar ratio of 30:1 for 1hr at room temperature. 1hr into stimulation, culture media was supplemented with 1X Brefeldin A (BioLegend) for the remaining 5hr of culture. Cells were then isolated, stained for cell viability using Live/Dead Fixable Staining Kit (Thermo Fisher) (1:60 dilution) as well as CD3 (clone OKT3), CD11c (clone S-HCL-3), CD14 (clone 63D3), CD16 (clone 3G8), CD56 (clone HCD56), and HLA-DR (clone L243) surface markers (BioLegend) (all at 1:30 diluton). After staining, cells were fixed and acquired using an Attune NxT flow cytometer (Invitrogen). NK cells were defined as viable CD3^-^CD14^-^CD16^+^CD56^+^HLA-DR^-^ cells (Supplementary Figure 1). NK cell degranulation was measured and reported as the fold change of NK cells positive for CD107a over control.

### Monocyte stimulation and cytokine measurements

Monocytes were isolated from healthy donor blood (Stanford Blood Center) using RosetteSep Human Monocyte Enrichment Kit (STEMCELL) per manufacturer instructions. Monocytes were cultured at a density of 2×10^6^ cells/mL in RPMI 1640 media supplemented with 1X non-essential amino acids, sodium pyruvate, penicillin-streptomycin-glutamine (Gibco), and 10% fetal bovine serum (GE Healthcare Life Sciences). Immune complexes were formed by incubating a dilution series of COVID-19 patient IgGs or anti-spike 3022 mAbs to SARS-CoV-2 spike-expressing delta-G-VSV pseudovirus for 1 hour at room temperature. Monocytes were incubated with the various immune complexes or the pseudovirus only for 18 hours at 37°C in a 5% CO_2_ incubator. After 18 hours, cell-free supernatants were collected and proinflammatory cytokine concentrations were measured using a LEGENDplex bead array (BioLegend) per manufacturer instructions.

### Statistical analysis

R statistical package version 1.2.1335 was used to generate the radar plots and perform multivariate linear regression analysis. All other data were analyzed with GraphPad Prism 8.0 software. Investigators were blinded to study subjects diagnoses during screening; COVID-19 patients and children were not known by investigators at the time of ELISA screening for RBD reactivity of serum or by investigators involved in relative quantitation of Fc glycoforms and IgG subclasses by mass spectrometry.

## Data availability

All raw data are available from the corresponding author on reasonable request. Reference SARS-CoV-2 spike protein sequence was obtained from the NCBI database (https://www.ncbi.nlm.nih.gov/protein/1791269090).

