## Supplementary Information for "Proinflammatory IgG Fc structures in patients with severe COVID-19"

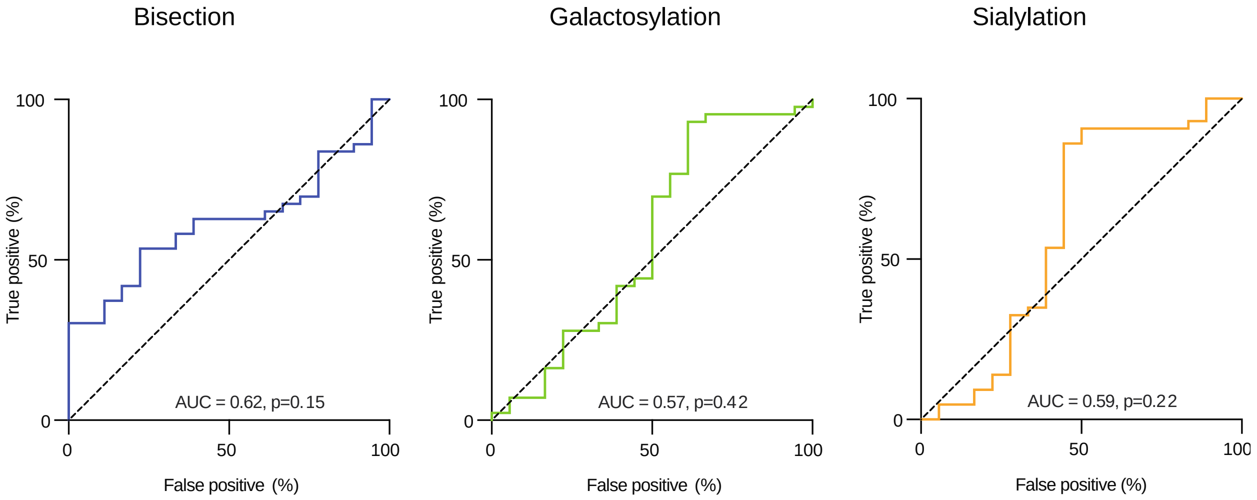


**Extended data Figure 1**: Receiver operating characteristic (ROC) curve for anti-RBD IgG1 IgG1 bisection (blue), galactosylation (green) and sialylation (yellow) from all hospitalized (n=43) and mild (n=18) COVID-19 patients showed these glycans were not predictors of COVID-19 severity. Area under the curve (AUC) for bisection 0.54[95% CI (0.38-0.69, p= 0.6357), galactosylation 0.61[0.42-0.81, p=0.1872 and sialylation 0.60[ 95% CI (0.40-0.79, p= 0.2245).

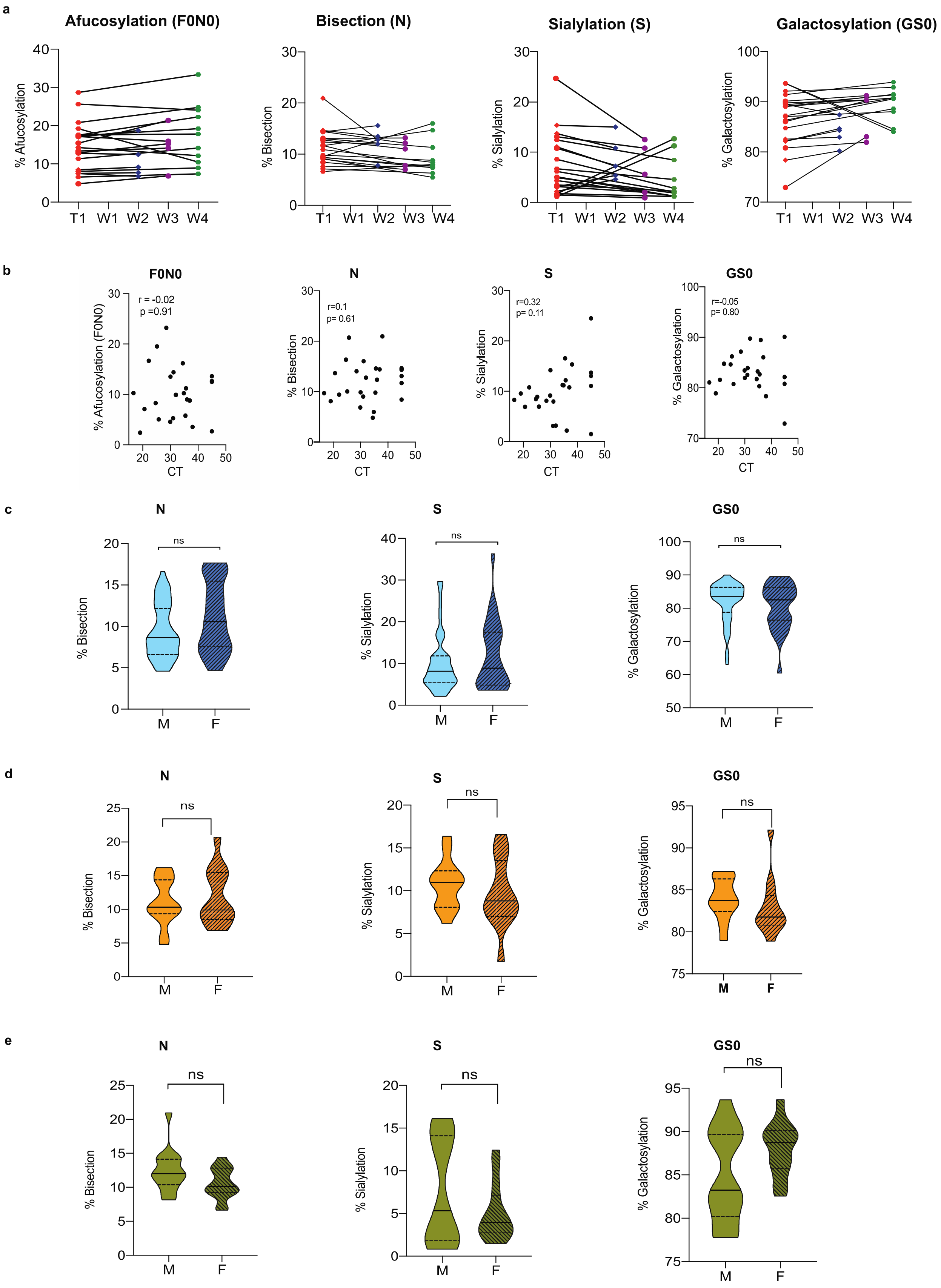
a
Afucosylation (F0N0)
Bisection (N)
Sialylation (S)
Galactosylation (GS0)
% Afucosylation
% Bisection
% Sialylation
% Galactosylation
T1 W1 W2 W3 W4
b
F0N0
N
S
GS0
r = -0.02
p =0.91
r=0.1
p= 0.61
r=0.32
p= 0.11
r=-0.05
p= 0.80
% Afucosylation (F0N0)
CT
c
N
S
GS0
ns
% Bisection
% Sialylation
% Galactosylation
M F
d
e

**Extended data Figure 2**: **(a)** Longitudinal analysis of the levels of anti-RBD IgG1 Fc afucosylation from COVID-19 patients at various time points. T1 represents the first draw with a second time point at week 2 (n=5), week 3 (n=5) or week 4 (n=11). Anti-RBD IgG1 from COVID-19 patients were characterized for Fc afucosylation (F0N0), bisection (N), sialylation (S) and galactosylation (GS0). None of the glycan levels were significantly different between T1 and subsequent time-points post infection. P values were calculated using two-tailed Wilcoxon matched-pairs signed rank test. **(b)** There was no correlation (Pearson's correlation coefficient (r) was calculated, and two-tailed p and r values have been reported) between the viral load and the abundance of any of the glycans on anti-RBD IgG1 (n=25). **(c, d)** The levels of the various glycans were not significantly different amongst male and female hospitalized COVID-19 patients in two cohorts (Blue-Kaiser Permanente (n=81, M=55, F=26), Orange-Stanford hospital (n=30, F=14, M=16)) or in **(e)** mild COVID-19 patients (n=27, F=14, M=13). Violin plots in (c), (d) and (e) show the distribution of sample values along with median (solid lines) and quartile (broken lines) values. P values were calculated using a two-tailed unpaired t test with Welch's correction.

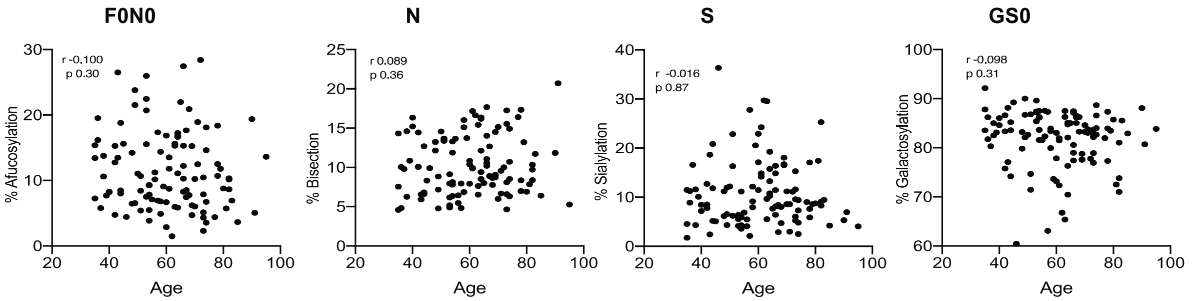


**Extended data Figure 3**: Age of COVID-19 patients (n=107) did not correlate with abundance of any of the Fc glycoforms. Pearson's correlation coefficient (r) and two-tailed p values have been reported.

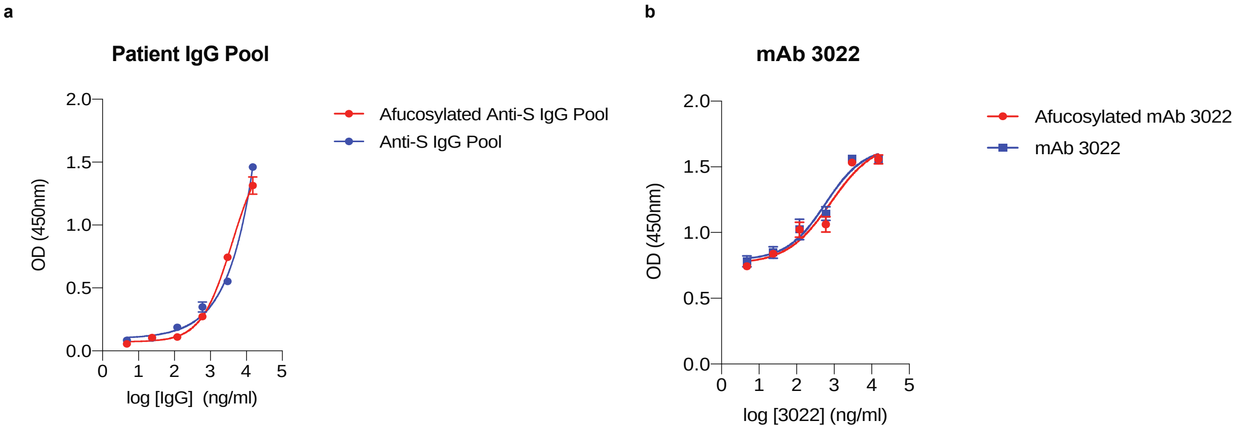


**Extended data Figure 4:** The level of IgG afucosylation did not impact binding to spike protein both for (a) pooled IgG from patients or (b) mAb 3022. The assays were performed in duplicate and mean data and standard deviation (SD) have been graphed.

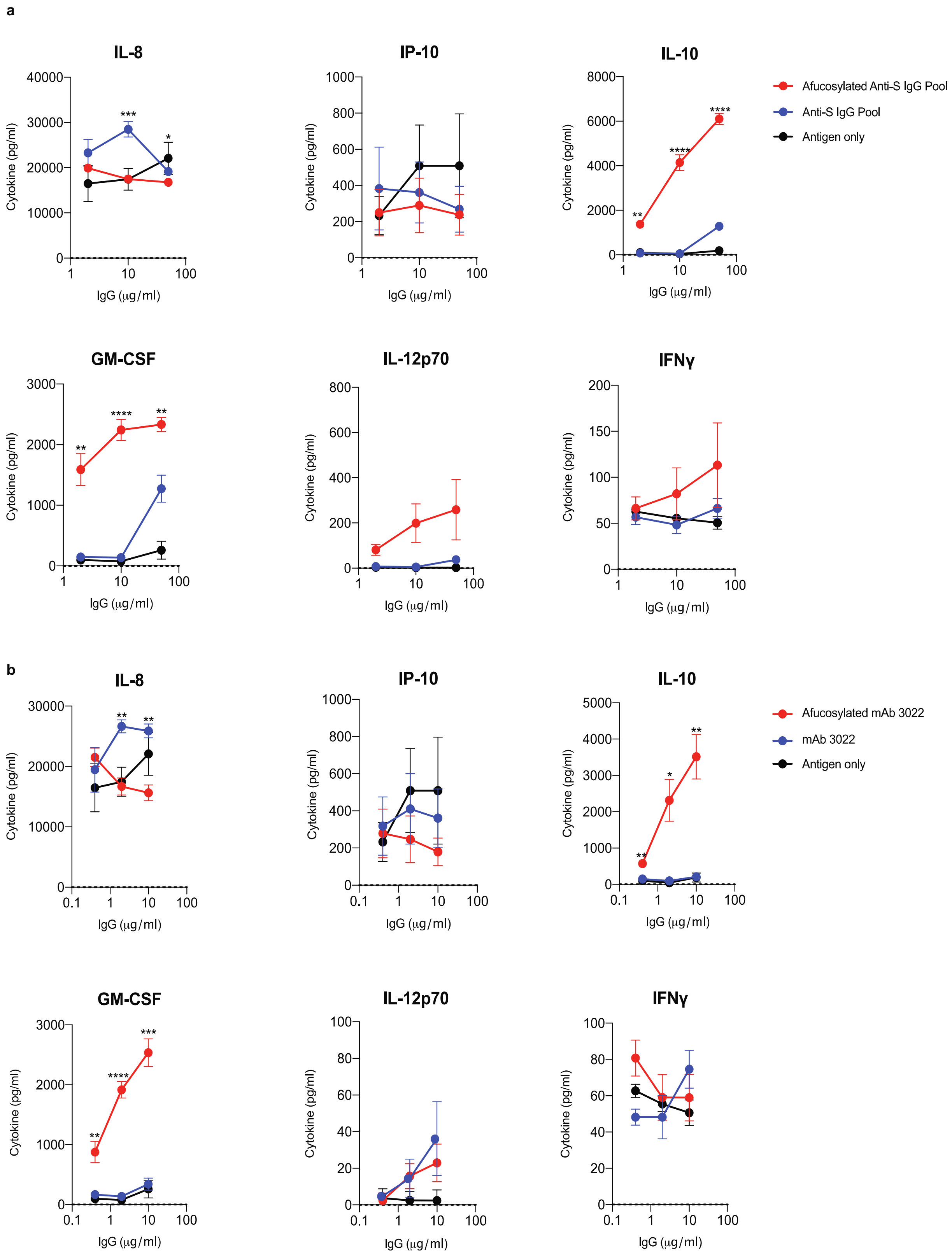


**Extended data Figure 5:** Monocytes stimulated with highly afucosylated immune complexes made from both **(a)** patient IgG pools (IL8: $p=0.0196$ for 100µg/ml, $p=0.0002$ for 20µg/ml. IL10: $p<0.0001$ for 100µg/ml, $p<0.0001$ for 20µg/ml and $p= 0.0019$ for 4µg/ml. GM-CSF: $p=0.0036$ for 100µg/ml, $p<0.0001$ for 20µg/ml and $p=0.003$ for 4µg/ml)

and **(b)** mAb 3022 (IL8: p=0.001 for 10µg/ml, p=0.0011 for 2µg/ml. IL10: p= 0.0045 for 10µg/ml, p=0.0112 for 2µg/ml and p= 0.0021 for 0.4µg/ml. GM-CSF: p=0.0007 for 10µg/ml, p<0.0001 for 2µg/ml and p=0.0036 for 0.4µg/ml) produced higher amounts of multiple proinflammatory cytokines as compared to immune complexes with low afucosylation. The assays were performed in duplicate with monocytes from three healthy donors and mean data and standard error of the mean (SEM) has been graphed. P-values between high and low afucosylated immune complexes at each antibody concentration were calculated by two-tailed paired t-tests. *P ≤ 0.05, **P ≤0.01, ***P ≤ 0.001, ****P ≤ 0.0001.

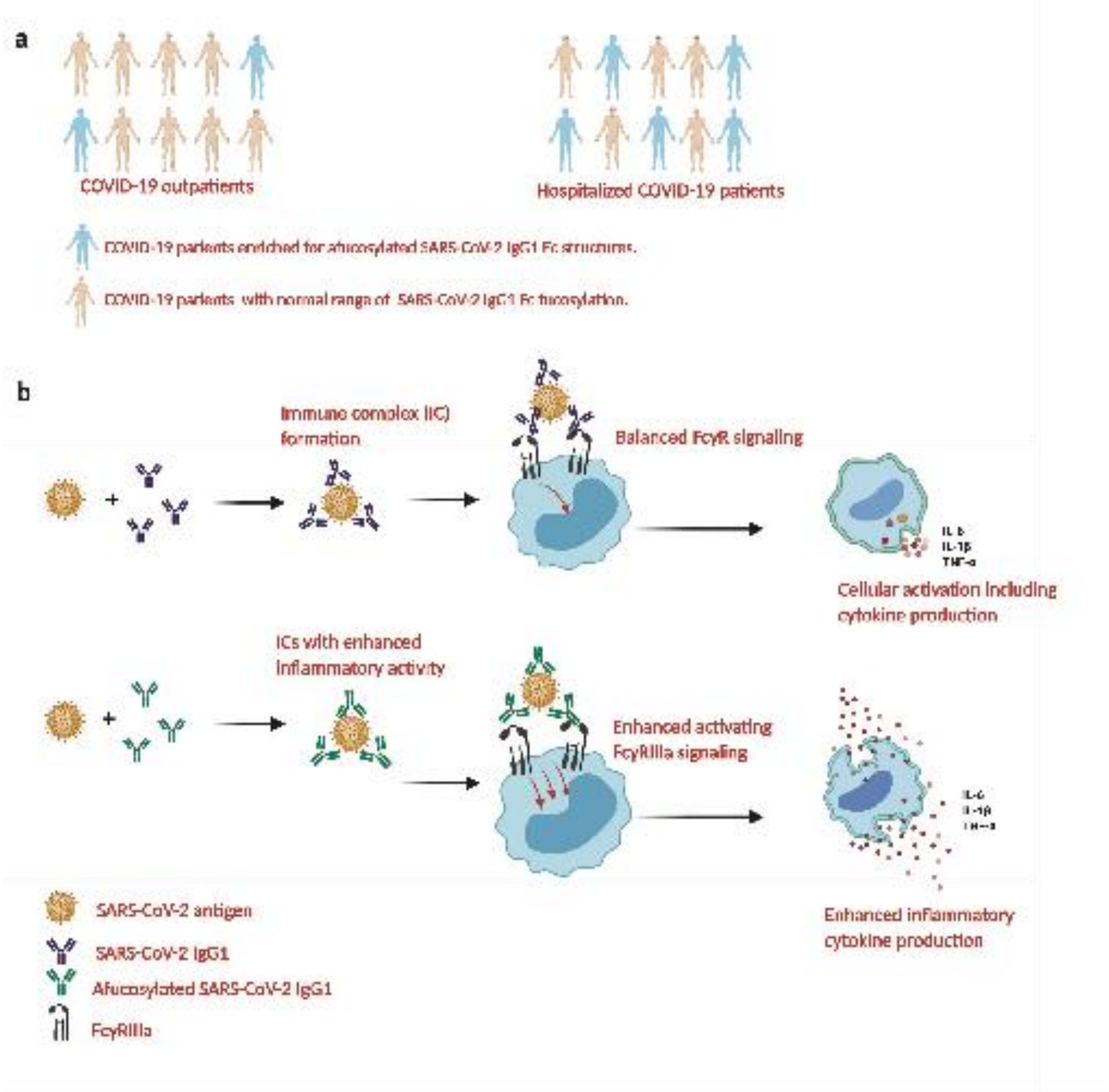


**Extended data Figure 6: (a)** Severe COVID-19 patients were more likely to have elevated levels of afucosylated IgG1 as compared to patients with mild disease. **(b)** Immune complexes formed from these high afucosylated antibodies have stronger binding to low affinity activating FcγRIIIa on surface of innate immune cells and thus more ITAM signaling. This leads to an production of pro-inflammatory cytokines.
